# ICI-induced Granulomatous Sialadenitis is Responsive to Prednisone

**DOI:** 10.64898/2026.01.21.26344113

**Authors:** Rachel J. Kulchar, Amara Ogbonnaya-Whittlesey, Margaret E. Beach, Zohreh Khavandgar, Eileen Pelayo, David E. Kleiner, Paola Perez, Daniel Martin, Peter Burbelo, John A. Chiorini, Paul Boutros, Douglas Wilmot Ball, Karim Boudadi, Maria Cabanillasis, Alan N. Baer, Blake M. Warner

## Abstract

Immune checkpoint inhibitors (ICIs) have transformed cancer treatment but commonly cause immune-related adverse events (irAEs), whether administered as monotherapy or in combination with other oncological agents. We present the first reported case of ICI-induced granulomatous sialadenitis in a male patient in his mid-fifties with BRAF-V600E-mutated papillary thyroid carcinoma who received sequential treatment with BRAF/MEK inhibitors followed by pembrolizumab. The patient experienced acute-onset severe xerostomia and salivary hypofunction, prompting ICI cessation and salivary gland biopsy. Integrative analysis using histology, single-cell RNA sequencing, and spatial transcriptomics revealed macrophage- and T-cell-mediated epithelial damage driven by epithelial senescence and Th1-polarized inflammation. Corticosteroid therapy reduced granuloma burden and improved salivary flow rates and tissue architecture; however, extensive fibrosis persisted despite treatment. These findings underscore the critical importance of early irAE recognition and intervention to preserve glandular function and enable continuation of cancer therapy.

## Introduction

Immune checkpoint inhibitors (ICIs) restore antitumor immunity by blocking inhibitory PD-1/PD-L1 interactions, enabling cytotoxic T cell-mediated tumor rejection (Postow et al., 2018). To enhance therapeutic efficacy, ICIs are frequently combined with CTLA-4 inhibitors, chemotherapies, or mutation targeted small molecule inhibitors such as BRAF and MEK inhibitors (Hamidi et al., 2024). However, these therapies increase the risk of immune-related adverse events (irAEs) affecting multiple organs, including the salivary glands, where clinical sialadenitis occurs in 5-35% of patients (Warner et al., 2019; Rosenberg et al., 2025). Due to limited epithelial regenerative capacity, ICI-induced inflammatory sialadenitis can progress to chronic sicca syndrome (ICI-S) characterized by persistent salivary hypofunction and xerostomia (Warner & Baer, 2021). These complications increase susceptibility to oral infections, impair quality of life, and may necessitate ICI discontinuation despite ongoing cancer treatment needs. While corticosteroids provide temporary symptom relief, disease-modifying therapies remain elusive (Warner & Baer, 2021).

Here we report the first case of ICI-associated granulomatous sialadenitis (ICI-GS) in a patient with BRAF-V600E-mutated papillary thyroid carcinoma treated sequentially with BRAF/MEK inhibitors followed by pembrolizumab. Granulomatous inflammation represents a distinct immunological response involving organized epithelioid macrophage and T cell aggregates; this histological presentation has not been previously documented as an ICI complication in salivary tissues, though it has been reported that BRAF/MEK inhibitors can induce sarcoid-like granulomas in skin and lung (Pham et al., 2022). We hypothesized that BRAF/MEK therapy established subclinical tissue inflammation or altered immune regulation, which was subsequently amplified by PD-1 blockade to trigger symptomatic granulomatous sialadenitis. To elucidate disease mechanisms and therapeutic response, we analyzed minor salivary gland biopsies obtained at presentation and following a five-week corticosteroid taper using integrated histological, single-cell RNA sequencing, and spatial transcriptomic analyses.

### Clinical Presentation and Sudden Onset of Severe Oral Dryness

A man in his mid-fifties with metastatic, poorly differentiated papillary thyroid carcinoma (pT2N1bMx) underwent total thyroidectomy and radioactive iodine ablation (Figure 1A, Supplemental Figure 1A). Six months postoperatively, PET/CT imaging revealed lung and rib metastases, upstaging the disease to pT3b, pN1b, cM1. Molecular profiling identified BRAFV600E and TERT promoter mutations, prompting initiation of combination BRAF/MEK inhibitor therapy with dabrafenib plus trametinib. After nine months of stable disease with persistent residual metastases, pembrolizumab (200 mg intravenously every three weeks) was added to the regimen. Following three cycles of pembrolizumab, the patient developed severe, acute-onset xerostomia that significantly impaired eating and sleep (**Figure 1A, B**). Pembrolizumab was discontinued after the fourth infusion due to suspected immune-related salivary gland toxicity.

**Figure 1:**
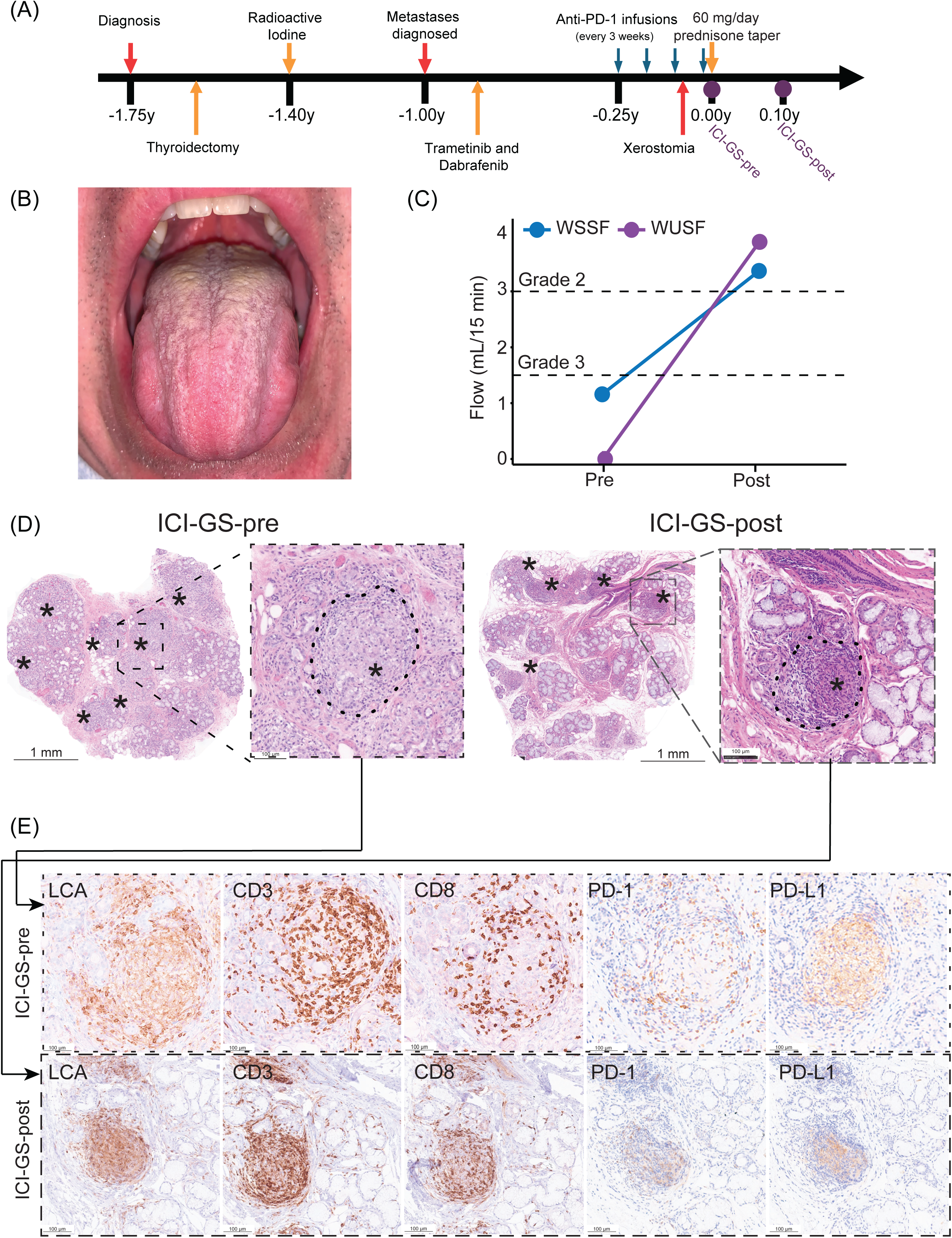
(A) Comprehensive clinical work up. The clinical timeline relative to when the patient was first evaluated by at the NIDCR Sjögren’s Disease Clinic at the NIH. The patient was diagnosed with papillary thyroid cancer ∼1.75 years before NIH evaluation. Metastatic disease was identified via PET CT scan 1 year prior to NIH evaluation. Treatment with trametinib (MEK1/MEK2 inhibitor) and dabrafenib (BRAF inhibitor) was initiated 0.9 years before NIH evaluation and continued throughout the prednisone taper. The first infusion of pembrolizumab was ∼0.25 years prior to NIH evaluation. These infusions continued every three weeks for a total of four infusions. The symptoms of dry mouth began after the third pembrolizumab infusion, around 0.1 years before NIH evaluation. The patient first visited the NIH 3 weeks after his fourth infusion of pembrolizumab (pre-treatment). After a 5-week steroid taper, the patient was evaluated again by the NIH (post-treatment). **(B) Clinical picture of patient’s oral cavity at follow-up post prednisone taper.** The image demonstrates a dry-appearing mucosa and coated tongue (keratin accumulation) that does not rub off. Examination of the patient’s oral cavity demonstrated remarkably improved oral status with reduced erythema and increased oral wetness. Within days of prednisone initiation, the patient reported being able to eat normally and resolution of joint pain. **(C) Whole unstimulated salivary flow (WUSF) and whole stimulated salivary flow rates (WSSF) rates (mL/15 min) in the patient with ICI-GS.** Flow was measured before (pre) and after (post) prednisone therapy. Dotted lines indicate salivary flow thresholds corresponding to objective CTCAE V6.0 Grades 2 and 3 salivary hypofunction for whole unstimulated salivary flow rates. **(D) Prednisone therapy treats granulomatous sialadenitis induced by checkpoint inhibitor therapy.** Histological examination (pathologist review: DEK, BMW) of hematoxylin and eosin (H&E) stained formalin-fixed, paraffin-embedded (FFPE) sections of the patient’s initial salivary gland biopsy revealed poorly organized non-necrotizing granulomas (*asterisks*) composed of epithelioid macrophages and a brisk lymphocytic infiltrate. After completion of the prednisone taper, a second salivary gland biopsy demonstrated a clinically important reduction in inflammation scores (focus score from 7 foci per 4mm^2^ to 3 foci per 4mm^2^ of salivary parenchyma) after prednisone taper. Although the glands appeared less inflamed, they also appeared atrophic and fibrotic. After prednisone, granulomas were reduced in number, smaller, and more densely organized. The initial biopsy demonstrated 22 granulomas enumerated among the 10.4 mm^2^ parenchymal tissue evaluated and affected all the salivary glands submitted. Posttreatment, 10 granulomas were enumerated among the 15.0 mm^2^ of parenchymal tissue evaluated. **(E) Immunohistochemistry staining of the ICI-GS-pre and ICI-GS-post tissue.** Immunophenotyping of the granulomatous inflammation using anti-leukocyte common antigen (LCA), -CD3, -CD4, - CD8, -PD-1, and -PD-L1 antibodies highlights the presence and arrangement of T-lymphocytes (anti-CD3 and anti-CD4 or anti-CD8), epithelioid macrophages (anti-LCA, anti-CD4, anti-PD-L1) in the granulomas, and the arrangement relative to affected epithelial structures.

The patient was subsequently referred to the NIDCR Sjögren’s Disease Clinic for evaluation, where examination revealed complete absence of stimulated salivary flow (CTCAE Grade 3 hypofunction) (**Figure 1C**). Minor salivary gland biopsies were obtained at this initial visit (designated ICI-GS-pre).

### Clinical Evaluation and Histopathology

During BRAF/MEK inhibitor therapy, the patient reported new-onset arthralgias affecting the knees, feet, and ankles, but denied other systemic symptoms including cutaneous lesions, respiratory symptoms, or mucosal involvement. Laboratory evaluation revealed no biochemical evidence of systemic granulomatous disease (**Supplemental Table 1**).

Salivary gland histopathology demonstrated marked sialadenitis with immune-mediated epithelial injury and extensive, poorly organized non-necrotizing granulomas involving all of the submitted glands (**Figure 1D**). Notably, the granulomas lacked the characteristic dense fibrosis typical of sarcoidosis. Microbial stains (acid-fast bacilli, Grocott methenamine silver, and periodic acid-Schiff) were negative; polarized light microscopy revealed no foreign material (**Supplemental Figure 1B**).

The inflammatory pattern and cellular composition were consistent with ICI-induced granulomatous inflammation (**Figure 1D–E**; **Supplemental Figure 1B**). Immunohistochemical analysis revealed granulomas composed of epithelioid macrophages (LCA⁺, CD68⁺) admixed with approximately equal numbers of CD4⁺ and CD8⁺ T cells (CD3⁺). PD-1⁺ T cells were abundant within and surrounding granulomas, while PD-L1 expression was restricted to macrophages within granulomas and to ductal and acinar epithelium involved by granulomatous inflammation (**Figure 1D–E**). B cells (CD20⁺) and plasma cells (CD138⁺) were notably absent from the inflammatory infiltrate.

### Clinical and Histological Response to Prednisone

The patient was treated with oral prednisone initiated at 60 mg daily and tapered over five weeks. By day 36 of treatment, the patient reported subjective improvement in xerostomia with restoration of the ability to eat solid foods without liquid assistance. Musculoskeletal pain also improved. Objective salivary function testing demonstrated marked improvement: unstimulated whole saliva flow increased from 0.0 to 3.6 mL/15 min (normal range: >1.5 mL/15 min), and stimulated flow increased from 1.2 to 3.3 mL/15 min (normal range: >7.5 mL/15 min) (**Figure 1C**). Follow-up labial salivary gland biopsy obtained at day 36 (ICI-GS-post) demonstrated appreciable reduction in granuloma burden, with overall more organized granulomas with mean granuloma density decreasing from 2.2 to 0.7 per mm² and individual granuloma size reduced. The overall inflammatory infiltrate was markedly decreased with fewer epithelioid macrophages. T cells remained the predominant lymphocyte population within residual granulomas, while CD20⁺ B cells remained sparse. Although both CD4⁺ and CD8⁺ T cells persisted within granulomas, the infiltrate shifted from a balanced CD4:CD8 ratio at baseline to the expected CD4⁺ T cell predominance post-treatment. PD-1 expression remained localized to T cells within granulomas, while PD-L1 expression was reduced and restricted to epithelioid macrophages (**Figure 1D–E**). Despite the reduction in acute inflammation, histological examination revealed significant epithelial parenchymal atrophy with prominent inter- and intra-lobular fibrosis, indicating incomplete tissue recovery and residual scarring.

## Results

### Immune-Engaged Epithelia and Inflammation Characterize ICI-GS and Resolve with Prednisone

Single-cell RNA sequencing (scRNA-seq) of salivary gland tissue obtained before and after prednisone treatment corroborated the histological findings and quantified cellular changes induced by ICI therapy and subsequent corticosteroid intervention (**Figure 2A**). Following quality control, dimensionality reduction, unsupervised clustering, and cell type annotation using established markers (**Supplemental Figure 1C**), cellular composition was compared across treatment conditions (**Figure 2B**).

**Figure 2:**
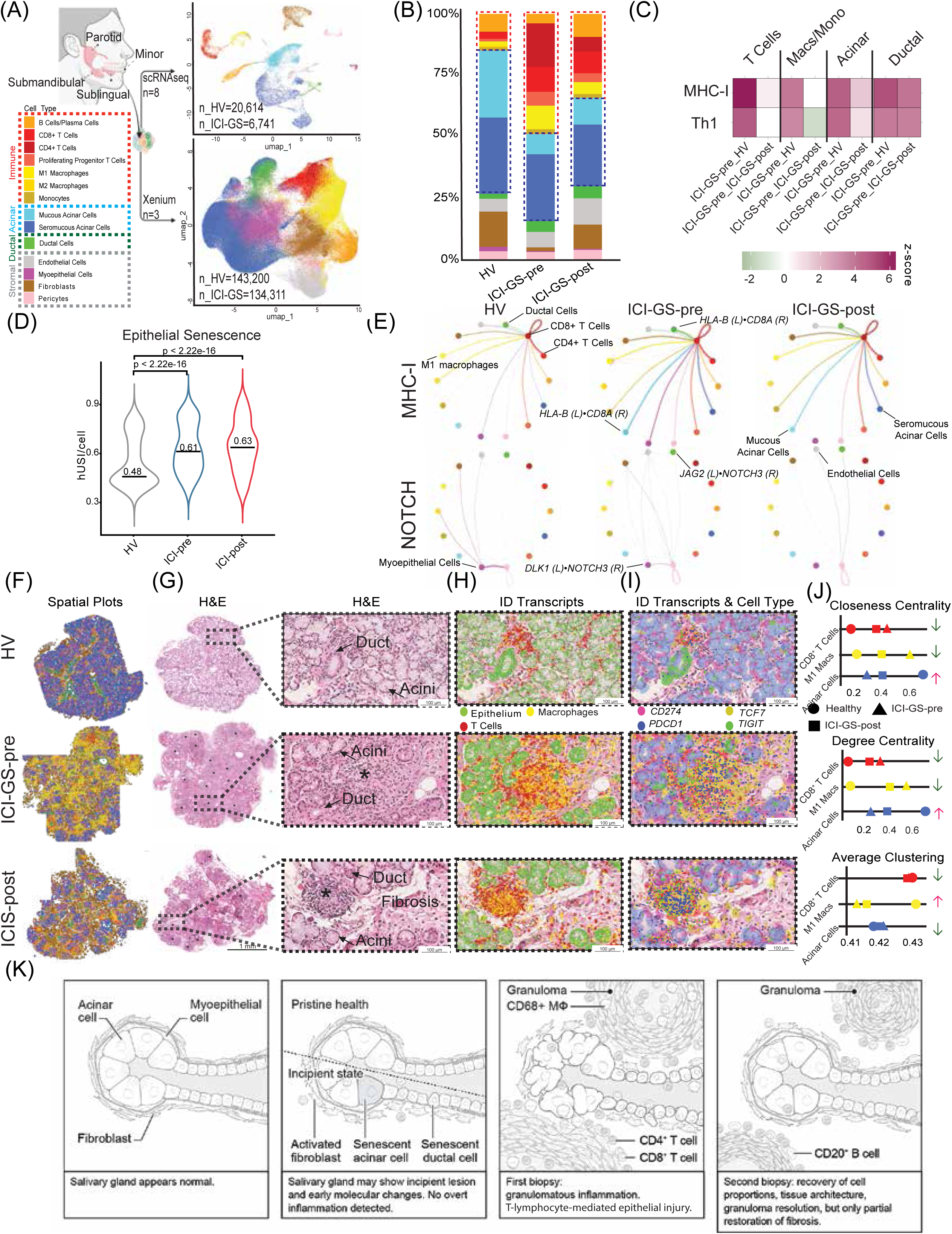
(A) Schematic overview of the methodology of this study and the integration of single cell and spatial transcriptomics to understand ICI-GS. Annotated scRNAseq UMAPs from dissociated human salivary gland biopsies. Twenty-two cell types were identified from 34,694 genes expressed among 27,355 cells. The annotated Xenium UMAPs from human salivary glands included 14 distinct cell populations, comprising 277,511 cells. (B) Percentage of each cell type by study cohort. (C) Heatmap comparing enrichment of canonical pathways conserved between HVs, ICI-GS-pre, and ICI-GS-post, colored by *z*-scores. Shown pathway comparisons have a p-value < 0.05. Enrichment values for each subcluster are relative ICI-GS-pre, with the pink color indicating that activation is greater or green indicating lesser in ICI-GS-pre. (D) Violin plot of senescence index ‘*expression’* among epithelial cells, with the median expression indicated by the horizontal line and text. Epithelial cells include ductal and acinar cells. The mean senescence expression in epithelial cells for HVs, ICI-GS-pre, and ICI-GS-post is 0.48, 0.61, and 0.63, respectively. The black line represents the median expression; ICI-GS-pre SG show a significant increase in senescence index to a healthy state (\*\*\*\**p* < 0.0001). **(E) Ligand-Receptor (L-R) interactions for HV and ICI-GS-pre groups for key senescent markers.** The names of the communicating cell types and representative L-R pairs are overlaid on the plot. **(F) Visualization of a salivary gland tissues across the three conditions**. **(G) H&E staining of the salivary glands by group.** The asterisks on the H&E scans indicate the presence of granulomas. **(H) H&E scans overlaid with gene markers for transcript location from spatial transcriptomics.** The green, red, and yellow dots represent composite epithelial, T cell, and macrophage gene markers, respectively. Visualization reveals enveloped epithelial cells within immune populations (e.g., macrophages) in the ICI-GS-pre tissue, which are almost entirely absent from both the tissues from a healthy patient and the ICI-GS-post. (I) Overlay of the H&E with cell type masks (coloring indicated in “Cell Type” legend [B]) and transcripts. *PD-1* and *PD-L1* are represented as *pink* and *blue* markers, respectively. Th1 genes, namely *TCF7* (*green* marker) and *TIGIT* (*yellow* marker) are also presented. **(J) Spatial analyses of subpopulations of cells within the tissue network by cell type and condition by closeness centrality, degree centrality, and average clustering.** ICI-GS-pre is represented as a triangle, ICI-GS-post as a square, and HVs as circles. Arrows up (red) or down (green) represent the change spatial architecture before and after prednisone, relative to HV tissues. The remaining list of clustering network analyses is provided in **SI Figure 1F. (K) Summary graphical vignette of the progression of granulomatous disease in the salivary glands compared to health and its response to prednisone therapy.**

The pre-treatment biopsy (ICI-GS-pre) demonstrated marked immune cell expansion compared to healthy control salivary glands. Total T cells increased from 4% to 34% of total cells (z-score = 10, p<0.001), *CD8⁺* cytotoxic T cells from 3% to 10% (z = 3, p<0.01), and M1-polarized macrophages from 2% to 10% (z = 5, p<0.001). Conversely, acinar cells, which comprise the functional secretory parenchyma, were significantly depleted from 58% in controls to 20% in ICIGS-pre (z = −2, p<0.05). Notably, a distinct population of immune-engaged (IE) acinar cells emerged, representing 15% of the total acinar compartment in ICI-GS-pre. These IE acinar cells exhibited gene expression signatures consistent with active T-cell-mediated targeting, including upregulation of antigen presentation machinery, interferon-stimulated genes, and stress response pathways.

Following the five-week prednisone taper, the post-treatment biopsy (ICI-GS-post) revealed partial normalization of cellular composition, with immune populations declining toward healthy proportions. Total T cells decreased from 34% to 18% (p<0.0001), *CD8⁺* T cells from 10% to 9% (p=0.1), and M1-polarized macrophages from 10% to 5% (p<0.0001) (**Figure 2B**). The acinar compartment showed modest recovery, with non-IE acinar cells increasing from 20% to 24% of total cells (p<0.0001), while IE acinar cells declined from 15% to 12% of the acinar population (p<0.0001), indicating partial but incomplete resolution of epithelial immune engagement.

### Epithelial Senescence and Immune Signaling Mark ICI-GS and Diminish with Corticosteroid Therapy

Recent evidence suggests that irAE pathogenesis involves T cell-mediated induction of epithelial senescence (Pringle et al., 2020). Consistent with this mechanism, IE acinar cells in ICIGS-pre glands exhibited broad upregulation of immune response pathways coupled with marked downregulation of genes essential for secretory function, including mucin genes (*MUC7*; 0.6-fold, p<0.0001) and calcium binding proteins (*ODAM*; 0.7-fold, p<0.0001) (**Figure 2C**). Following corticosteroid therapy, immune pathway activation was attenuated and secretory gene expression partially restored, paralleling the patient’s clinical improvement in salivary function.

To quantify epithelial senescence, we calculated a composite senescence index based on expression of established senescence-associated genes (Wang et al., 2025). The senescence index was significantly elevated in ICI-GS-pre acinar cells compared to healthy controls (0.61 vs. 0.48, p<0.00001), driven by increased expression of canonical senescence markers including *CDKN2A* (p16^INK4a^), and senescence-associated secretory phenotype (SASP) components *IFNG* and *IL1B* (**Figure 2D**). Notably, despite reductions in inflammatory cell infiltration, the epithelial senescence index remained persistently elevated following prednisone treatment (ICI-GS-post: 0.63 vs. ICI-GS-pre: 0.61, p=0.42), indicating that corticosteroids suppressed immune inflammation and partially restored canonical secretory function without reversing epithelial senescence.

Ligand-receptor interaction analysis revealed extensive immune-epithelial crosstalk mediated by key inflammatory pathways including TNF, IFNG, and NOTCH signaling (**Figure 2E**). MHC class I-mediated interactions between epithelial cells and *CD8⁺* T cells were markedly amplified in ICI-GS-pre and declined following corticosteroid therapy, consistent with reduced *CD8⁺*T cell infiltration. Collectively, these findings implicate epithelial senescence and sustained immune-epithelial interactions as central mechanisms driving granuloma formation and tissue damage in ICI-GS. The novel granulomatous presentation and incomplete therapeutic response prompted further investigation using spatial transcriptomics to define tissue architecture and cellular interactions *in situ*.

### Spatial Profiling Reveals Immune-Mediated Architectural Collapse and Recovery Following Prednisone Treatment

Spatial transcriptomic analysis of ICI-GS tissue revealed profound alterations in cellular composition and spatial organization compared with healthy salivary glands (**Figures 2F**, **Supplemental Figure 1D,E**). Pre-treatment ICI-GS specimens exhibited complete loss of normal glandular architecture, with granulomatous lesions dominated by M1-polarized macrophages and T cells. Following prednisone therapy, granulomas decreased markedly in both size and frequency (**Figure 1D**, **Figures 2F–I**).

Integration of spatial transcriptomic data with matched histology revealed that granulomas showed polarization toward epithelial structures such as ducts and acini which showed histological and transcriptomic evidence of immune-mediated injury. M1-polarized macrophages within pre-treatment granulomas co-expressed epithelial markers, indicating active phagocytosis of dying glandular epithelium (**Figures 2G–I**). This finding provides direct molecular evidence for immune-mediated destruction of secretory tissue in untreated ICI-GS.

Post-prednisone tissue demonstrated substantial reduction in overall immune infiltration, with decreased abundance of M1-polarized macrophages and T cells, accompanied by partial restoration of epithelial and stromal architecture (**Figure 2B**, **Supplemental Figure 1D**). However, immune cell populations remained elevated relative to healthy salivary gland controls, indicating incomplete immunological resolution. The post-treatment landscape was further characterized by increased fibroblast accumulation and expansion of B cell and plasma cell populations following prednisone taper, consistent with fibrotic remodeling and partial normalization of immune homeostasis (**Figure 1D**, **Figures 2C,D**).

Supporting these results, spatial neighborhood analysis of cellular interactions revealed re-establishment of physiological cell-cell associations among fibroblasts, acinar cells, ductal cells, and B-lymphocytes/plasma cells in post-treatment tissue (**Figures 2I,J**; **Supplemental Figures 1E,F**). These interaction patterns demonstrate partial restoration of tissue architecture and reduced immune-mediated injury. However, the combination of incomplete immunological resolution and sustained fibrotic remodeling likely maintains residual epithelial cells in a senescent state, potentially limiting functional recovery despite this architectural reorganization.

### BRAF/MEK-Induced Senescence Primes Epithelium for T Cell Mediated Injury Following PD-1 Blockade

Integrated analysis of ICI-GS-pre salivary glands revealed Th1-polarized granulomatous inflammation (**Figure 2C**), supporting a two-hit pathogenic model: BRAF/MEK inhibitors induced epithelial senescence during nine months of therapy (evidenced by elevated senescence index and SASP gene expression), establishing a pro-inflammatory microenvironment that was subsequently amplified by PD-1 blockade, unleashing T cell-mediated inflammation resulting in granulomas (**Figure 2K**). This hypothesis is consistent with the patient’s clinical course of subclinical systemic inflammation (arthralgias) during BRAF/MEK therapy followed by acute onset severe sialadenitis within weeks of pembrolizumab initiation.

The granulomatous inflammation exhibited a sarcoidosis-like molecular signature characterized by significant upregulation of *NOD2* (nucleotide-binding oligomerization domain containing protein 2; *p*<0.001), a pattern recognition receptor that senses cellular stress and activates granulomatous responses. Downstream NF-κB pathway components were coordinately upregulated, including *NFKB1*, *NFKBIA*, and *RELA* (all *p*<0.001), suggesting that senescence-associated signals triggered NOD2-dependent granuloma formation (Murphy & Weaver, 2017).

Granuloma-associated macrophages demonstrated pronounced M1 polarization with elevated expression of pro-inflammatory markers: *CD86, IL1B*, *TNF,* and *NOS2* (all *p*<0.001). Both CD4⁺ and CD8⁺ T cells exhibited activated cytotoxic effector phenotypes with significantly elevated cytotoxic molecules including *PRF1* (perforin), *GZMB* (granzyme B), and *IFNG* (all p<0.001 vs. healthy controls).

Notably, *CD4*⁺ T cells displayed this cytotoxic phenotype alongside canonical Th1 polarization markers: *TBX21*, *STAT4*, and *IL12RB1/*2 (IL-12 receptors; all p<0.001). Cytotoxic *CD4⁺* T cells are uncommon and typically associated with chronic autoimmune conditions, where they can directly mediate epithelial damage. Both T cell populations demonstrated markedly elevated *PDCD1* (in both *CD4⁺* and *CD8⁺* T cells; p<0.001), indicating sustained activation despite pembrolizumab blockade and suggesting chronic antigen stimulation by senescent epithelial cells. These findings support epithelial senescence and Th1/cytotoxic immune responses as central pathogenic mechanisms in ICI-associated granulomatous sialadenitis.

## Discussion

### Epithelial Senescence and Immune Checkpoint Disruption Drive Granulomatous Inflammation

Our findings support a sequential two-hit model of pathogenesis. BRAF/MEK inhibitors induce cellular senescence and enhance antigen presentation (Frederick et al., 2013), and our data suggest similar effects in salivary epithelium. The elevated senescence index, upregulation of SASP components, and loss of secretory genes indicate that nine months of BRAF/MEK therapy established widespread epithelial senescence. During this period, immune checkpoints likely restrained inflammatory responses, as evidenced by subclinical presentation with only arthralgias.

Pembrolizumab disrupted this balance, disinhibiting Tc/Th1 responses against senescent epithelium. The inflammation was characterized by NOD2-NF-κB pathway activation and a predominance of cytotoxic T cells including CD4⁺ T cells, rare outside chronic autoimmune conditions, alongside CD8⁺ T cells. Ligand-receptor analysis revealed extensive immune-epithelial crosstalk mediated by TNF, IFNG, and MHC class I interactions, driving granuloma formation (Figure 2K). Critically, epithelial senescence persisted after corticosteroid therapy, indicating that steroids suppress inflammation without reversing underlying epithelial pathology highlighting future therapeutic directions.

### Corticosteroids Resolve Acute Inflammation but Fail to Prevent Fibrosis

Prednisone treatment produced substantial clinical improvements for the patient including improvements in salivation and reduced feelings of a dry mouth, the ability to chew and swallow foods without the need to drink water between bites, reduced arthralgias. After prednisone, the glands demonstrated pathological improvement: granuloma burden decreased 68%, inflammatory infiltrates declined (T cells: 34%→18%; M1 macrophages: 10%→5%). However, post-treatment biopsies revealed extensive fibrosis with acinar atrophy, and the acinar compartment remained severely depleted (24% vs. 58% in controls). This dissociation between inflammatory resolution and progressive fibrosis likely reflects persistent senescent epithelium serving as a continuous source of pro-fibrotic signals. Thus, corticosteroids alone are insufficient to prevent irreversible damage; early intervention combining anti-inflammatory therapy with senolytic and anti-fibrotic strategies may be necessary to preserve glandular function.

### Clinical Implications

At 28-month follow-up, the patient’s disease had progressed, requiring alternative therapies. Whether continued pembrolizumab would have provided durable disease control remains unknown. The irreversible glandular damage from delayed intervention eliminated the possibility of ICI rechallenge, representing a forced trade-off between quality of life and potentially life-prolonging therapy. This case emphasizes critical medical needs including: (1) heightened surveillance for early salivary symptoms in ICI-treated patients; (2) prompt corticosteroid intervention; (3) investigation of adjunctive senolytics/anti-fibrotics; and (4) biomarker development for risk stratification to preserve both cancer therapy options and quality of life.

## Conclusions

We report the first case of ICI-associated granulomatous sialadenitis, a severe immune-related toxicity arising from sequential BRAF/MEK inhibition and PD-1 blockade. Integrated single-cell, spatial, and histologic analyses reveal that BRAF/MEK inhibitors induced epithelial senescence, which was subsequently amplified by pembrolizumab, unleashing cytotoxic T cells and M1 macrophages that drove granuloma formation and epithelial destruction.

While corticosteroids reduced inflammation and partially restored salivary function, epithelial senescence persisted and fibrosis developed, indicating incomplete recovery despite clinical improvement. These findings suggest that corticosteroids alone are insufficient and that early intervention combining anti-inflammatory, senolytic, and anti-fibrotic strategies may be necessary to prevent irreversible glandular damage.

This case underscores a critical clinical challenge: severe irAEs can necessitate discontinuation of potentially life-prolonging cancer therapies. The patient’s disease progression following pembrolizumab cessation highlights the importance of early recognition and aggressive management of salivary gland toxicity to preserve both quality of life and therapeutic options. Clinicians should maintain heightened surveillance for salivary and oral irAEs in patients receiving ICIs, particularly following prior BRAF/MEK therapy.

Our experience demonstrates that integrated molecular approaches, combining single-cell transcriptomics, spatial transcriptomic and proteomic profiling, and detailed histopathology, can elucidate irAE mechanisms and guide personalized management strategies for these increasingly common and complex immune toxicities.

## Ethics approval and consent to participate

All patients, including the patient discussed in the case report, provided written informed consent to NIH Central IRB-approved research protocol (15-D-0051, PI-MB), including consent for clinical photographs for the purposes of this case report, before any study procedures were initiated.

## Consent for publication

All authors have read the manuscript and consent to its publication.

## Availability of data and material

All scripts and pipelines used for data processing and analysis are available at https://github.com/rachelkul/ICI-sicca. Code can be freely used and adapted with appropriate citation. The spatial transcriptomic datasets generated and analyzed during the current study are available in Mendeley Data under accession number doi: 10.17632/ndpj26z5nb.1. Proper citation is required at: Warner, et al., (2025), “Spatial Transcriptomics of Minor Salivary Glands in Salivary Gland Diseases”, Mendeley Data, V1, doi: 10.17632/ndpj26z5nb.1 and this manuscript. Single cell RNA sequencing data are available at dbGaP and will be available upon publication dbGaP Study Accession: phs002446

## Competing interests

BMW receives funding for research from Pfizer, Inc., AstellasBio, Inc., Sjögren’s Foundation, and the Foundation for the National Institutes of Health. The work was not supported or influenced by these funding mechanisms.

## Funding

This research was entirely supported by the Intramural Research Program of the National Institutes of Health (NIH) through grants and awards to BMW (NIH/NIDCR Division of Intramural Research Z01-DE000704). Additional contributions were provided through core support from the NIDCR Genomics and Computational Biology Core. The contributions of the NIH author(s) are considered Works of the United States Government. The findings and conclusions presented in this paper are those of the author(s) and do not necessarily reflect the views of the NIH or theU.S. Department of Health and Human Services.

## Author’s contributions

Original Conceptualization: PP, RJK, ANB, BMW; Data Curation: RJK, AOW, MEB, ZK, EP, DEK, PP, DM, PB, DWB, KB, MC, BMW; Formal Analysis: RJK, DEK, BMW; Funding Acquisition: BMW; Investigation: RJK, DEK, AOW, PP, DM, BMW; Methodology: RJK, PP, DM, JAC, PB, BMW; Project Administration: BMW; Resources: JAC, BMW; Software: RJK; Supervision: BMW; Validation: RJK, PP, DM; Visualization: RJK, PP, BMW; Writing – Original Draft Preparation: BMW, AOW, RJK; Writing – Review & Editing: RJK, PP, ANB, PB, BMW.

## Supporting information

Supplemental Table 1. Clinical testing results at initial and follow-up evaluations

Supplemental Table 2. Clinical characteristics of the single cell and spatial transcriptomic data

## Data Availability

All scripts and pipelines used for data processing and analysis are available at https://github.com/rachelkul/ICI-sicca. Code can be freely used and adapted with appropriate citation. The spatial transcriptomic datasets generated and analyzed during the current study are available in Mendeley Data under accession number doi: 10.17632/ndpj26z5nb.1. Proper citation is required at: Warner, et al., (2025), Spatial Transcriptomics of Minor Salivary Glands in Salivary Gland Diseases, Mendeley Data, V1, doi: 10.17632/ndpj26z5nb.1 and this manuscript. Single cell RNA sequencing data are available at dbGaP and will be available upon publication dbGaP Study Accession: phs002446

## Acknowledgements

The authors are deeply thankful to the patients, and the NIDCR Office of the Clinical Director, NIDCR Office of the Scientific Director, and the staff of the NIDCR Dental Clinic for their support and contributions. The authors also appreciate the helpful suggestions from Dr. Weixu Wang, cocreator of the Human Universal Senescence Index, in utilizing this tool to estimate senescence in our patient populations.

## Notes

### Competing Interest Statement

BMW receives funding for research from Pfizer, Inc., AstellasBio, Inc., Sjogren's Foundation, and the Foundation for the National Institutes of Health. The work was not supported or influenced by these funding mechanisms.

### Author Declarations

All patients, including the patient discussed in the case report, provided written informed consent to NIH Central IRB-approved research protocol (15-D-0051, PI-MB), including consent for publication of the case and clinical photographs for the use in this case report.

