## Supplemental Table 1. Clinical testing results at initial and follow-up evaluations for "ICI-induced Granulomatous Sialadenitis is Responsive to Prednisone"

### *Supplemental Table and Figure.*

| **Supplemental Table 1. Clinical testing results at initial and follow-up evaluations** | | | |
|---|---|---|---|
| Test name | Initial visit | Follow-up visit, 36 days after Initial visit. | Normal Range |
| ANA Hep-2 substrate S IgG[1] | ***Positive, 1:160*** | ***Positive, 1:160*** | <1:80 |
| ENA[2] | ***SSA 6.9 units***<br>SSB <0.2 units<br>Anti-RNP <0.2 units<br>Anti-Sm <0.2 units<br>Scl 70 <0.2 units<br>Anti-Jo-1 <0.2 units | ***SSA 5.9 units***<br>SSB <0.2 units<br>Anti-RNP <0.2 units<br>Anti-Sm <0.2 units<br>Scl 70 <0.2 units<br>Anti-Jo-1 <0.2 units | <1 units |
| Ro52-LIPS | ***$2.6*10^6$ RLU***[3] | ***$1.6*10^6$ RLU*** | $<10^4$ RLU |
| Ro60-LIPS | $<10^4$ RLU | $<10^4$ RLU | $<10^4$ RLU |
| Quantitative immunoglobulins | IgG 1668 mg/dL<br>IgA 410 mg/dL<br>IgM 89 mg/dL | IgG 1441 mg/dL<br>IgA 380 mg/dL<br>IgM 80 mg/dL | IgG 540-1822 mg/dL<br>IgA 63-484 mg/dL<br>IgM 22-240 mg/dL |
| C3 complement | 135 mg/dL | 126 mg/dL | 82-185 mg/dL |
| C4 complement | 35 mg/dL | 31 mg/dL | 15-53 mg/dL |
| RF[4] | <10 units | <10 units | <10 units |
| ACE[5] | 50.6 units | Not performed | 3-52 units/L |
| Schirmer test without anesthesia | ***Right eye 4mm/5 min***<br>***Left eye 3mm/5 min*** | ***Right eye 5mm/5 min***<br>Left eye 7mm/5min | Right eye 6 mm/5min<br>Left eye 6 mm/5min |
| Van Bijsterveld[6] | Right eye 1<br>Left eye 0 | Not performed | Right eye <4<br>Left eye <4 |
| Unstimulated whole saliva flow rate | ***0 mL/15 min*** | 3.60 mL/15 min | >3.0 mL/min |
| Minor salivary gland Focus score | 7 | 3 | 0 |

Bolded and italicized values indicate they fall outside the normal range. *Abbreviations.* Anti-nuclear antibody Hep-2 substrate S immunoglobulin G; 2. Extractable Nuclear Antigens; 3. Relative Light Units; 4. Rheumatoid factor; 5. Angiotensin converting enzyme; 6. Corneal staining.
