## Supplementary figures and images for "ICI-induced Granulomatous Sialadenitis is Responsive to Prednisone"

### Supplemental Figure 1.

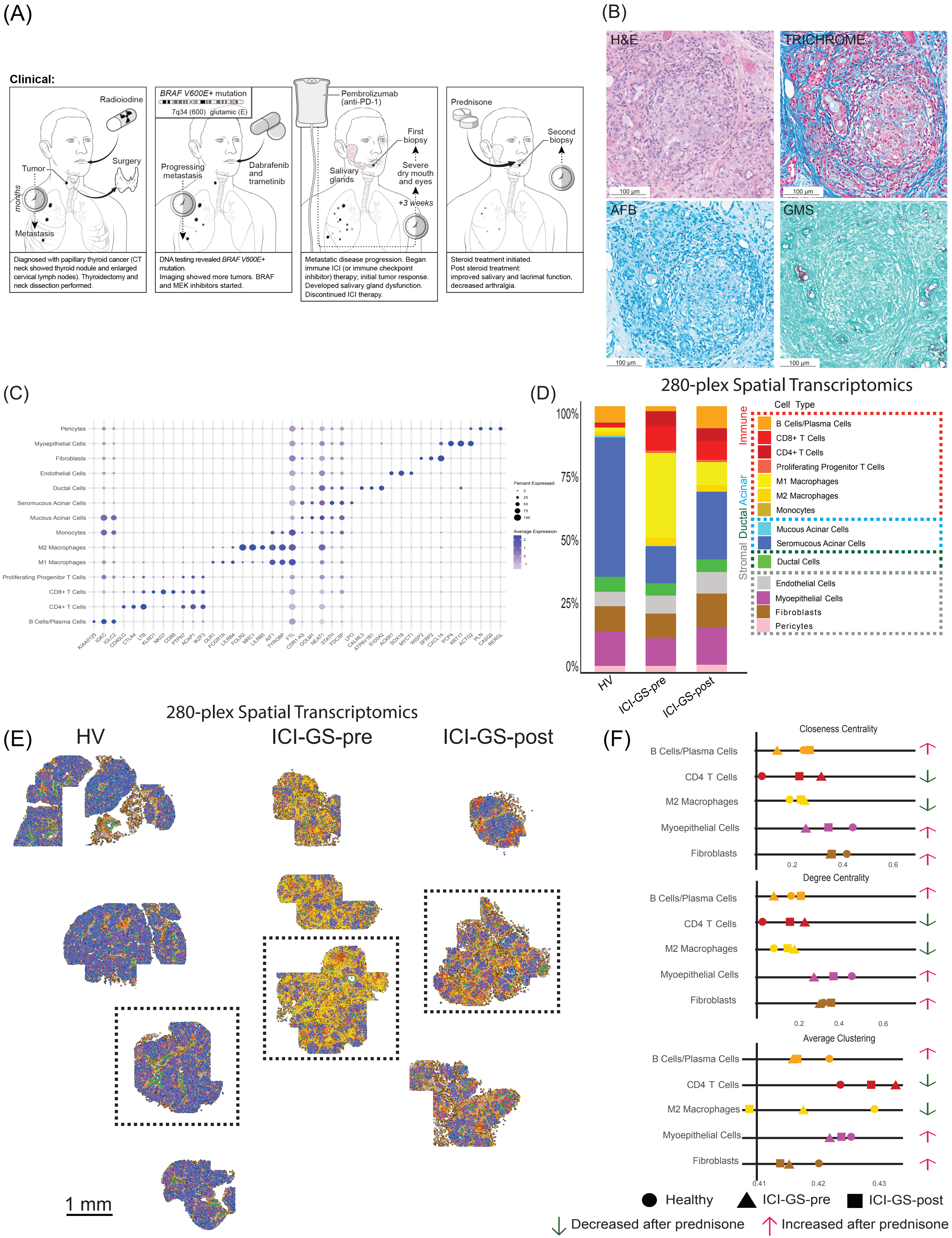
