## Supplemental Figure 1 Legend for "ICI-induced Granulomatous Sialadenitis is Responsive to Prednisone"

***Supplemental Figure Legends.***

**Supplemental Figure 1: (A) Clinical graphical abstract of the clinical chronology. (B) Histochemical staining of ICI-GS.** Stains for trichrome, Grocott's methenamine silver (GMS), and acid-fast bacteria (AFB) reveal the absence of microorganisms and polarizable foreign material in the granulomas. **(C) scRNAseq dot plot of the top 3 most differentially expressed genes by cluster.** The circles are colored by average expression levels, with the size referring to the percent of cells that express a certain gene by cluster. **(D) Proportion of cell types within the SGs by patient cohort for the Xenium data. (E) Whole slide spatial plots.** Tissues samples are from HV #1 and the patient with ICI-GS before and after prednisone treatment. **(F) Spatial closeness centrality, degree centrality, and average clustering networks plotted by condition and cluster.** ICI-GS-pre is represented as a triangle, ICI-GS-post as a square, and HVs as circles. Arrows up or down represent the change spatial architecture before and after prednisone, relative to HV tissues.
