## Supplemental Table 2. Clinical characteristics of the single cell and spatial transcriptomic data for "ICI-induced Granulomatous Sialadenitis is Responsive to Prednisone"

| Supplemental Table 2. Clinical characteristics of the single cell and spatial transcriptomic data | | | | | | |
|---|---|---|---|---|---|---|
| **Spit Number*** | **Sex** | **Race** | **Ethnicity** | **Histopathological Interpretation** | **Focus Score** | **Case** |
| 3717 | Female | White | Not Latino or Hispanic | Mild Chronic Sialadenitis | 0 | 0 |
| 3735 | Female | White | Latino or Hispanic | Normal | 0 | 0 |
| 3743 | Female | White | Not Latino or Hispanic | Mild Chronic Sialadenitis | 0 | 0 |
| 3632 | Female | White | Not Latino or Hispanic | Mild Chronic Sialadenitis | 0 | 0 |
| 3633 | Female | White | Not Latino or Hispanic | Mild Chronic Sialadenitis | 0 | 0 |
| 3437 | Female | Black or African American | Not Latino or Hispanic | Mild Chronic Sialadenitis | 0 | 0 |
| 3741_pre** | Male | White | Not Latino or Hispanic | Severe Chronic Sialadenitis | 7 | 1 |
| 3741_post** | Male | White | Not Latino or Hispanic | Mild Chronic Sialadenitis | 3 | 1 |

***** - Spit Numbers are unknown to anyone outside the research group.

**** -** Case had biopsies taken for clinical purposes before (“pre”) and after (“post”) effective prednisone taper.
