## Appendix ∼ Supplemental Information for "ICI-induced Granulomatous Sialadenitis is Responsive to Prednisone"

*ICI-GS Patient Information*

The clinical timeline of the patient's diagnosis and treatment plan is provided in **Figure 1A** and **Supplemental Figure 1A**. The patient's past medical history was notable for wellcontrolled hypertension, managed with an angiotensin receptor blocker. He was a non-smoker and drank 5-7 standard alcoholic drinks per week. He had a sibling with possible psoriasis and arthritis but no other relevant family history.

The patient's cancer was first diagnosed 20 months before visiting our clinic during an evaluation for left-sided cervical lymphadenopathy and a 2.6 centimeter (cm) left lobe thyroid nodule. Fine needle aspiration biopsy of the thyroid nodule and a level 3 cervical node demonstrated papillary thyroid carcinoma. The patient underwent total thyroidectomy, cervical neck dissection, and excision of a retropharyngeal 1.6 cm mass at the skull base. Next generation sequencing of the tumor showed *BRAF V600E* and *TERT* promoter mutations. The surgery was followed by radioiodine ablation using an oral dose of 30 mCi. Post-treatment, there were three foci of radioiodine uptake confined to the thyroid bed on whole body planar I123 scan and an undetectable stimulated thyroglobulin level. Lung nodules decreased from 17 millimeters (mm) to 4 mm or less; however, a thyrogen-stimulated whole-body scan showed no focal uptake, precluding further radioiodine treatment. Six months after the original cancer diagnosis, PET-CT scanning showed progressive disease with FDG-avid lesions centered in the perihilar regions of both lungs, cervical nodes, and the thyroid surgical bed.

The patient had started co-therapy with dabrafenib and trametinib 11 months before visiting our clinic. Papilledema occurred as a rare side effect, resolved when the drugs were temporarily held, and did not recur when they were resumed. Pembrolizumab was started 18 months after cancer diagnosis and 12 weeks before his evaluation in our clinic. After the first dose of pembrolizumab, the patient developed an acneiform eruption of his central face and midline back and persistent pain in his knees and Achilles tendons. The patient denied ocular dryness. Ocular surface staining was absent when assessed with fluorescein and lissamine green dyes. Schirmer test without anesthesia showed 4 mm/5 min wetting in the right eye and 3 mm/5 min in the left eye, reflecting markedly reduced tear flow in both eyes. Salivary gland ultrasound showed mildly abnormal echogenicity and homogeneity of both parotid glands. On examination, there was lingual papillary atrophy and erythema, absent sublingual salivary pooling, angular cheilitis, rhinophyma, and an erythematous scaly rash confined to the eyebrows. At initial visit, the patient was diagnosed with oral candidiasis and fluconazole therapy was initiated.

Laboratory testing showed his white blood count (WBC) was 4850/mm$^3$, hemoglobin 13.6 g/dL, platelet count 190,000/mm$^3$, ESR 38 mm/hr, IgG 1668 mg/dL, and IgG4 31.5 mg/dL. There was 1+ proteinuria. Angiotensin-converting enzyme (ACE) level was 51 U/L. ANA was 1:160 with nucleolar staining, anti-SSA antibodies were 5.9 ELISA units (normal <1.0)

(**Supplemental Table 1**); ANA or SSA values prior to starting pembrolizumab were not available. Rheumatoid factor was negative. High levels of anti-Ro52 (2.6 x106. RLU), but not anti-Ro60 antibodies, were detected using a luciferase immunoprecipitation system (LIPS) assay.[1]

Our treatment approach included a prednisone taper--which was initiated as 60 mg daily for one week, 40 mg daily for one week, 20 mg daily for one week, and 10 mg daily for 2 weeks, followed by its cessation--along with oral fluconazole for oral candidiasis. After 5-week taper, Schirmer tests without anesthesia showed tear flow improvement (5 mm/5 min wetting in the right eye and 7 mm/5 min in the left eye).

The month following final evaluation at the National Institutes of Dental and Craniofacial Research's Sjögren's Disease Clinic, the patient discontinued Dabrafenib (December 2022). In August 2023, he also discontinued trametinib and began binimetinib plus encorafenib.

*Tissue Dissociation of Human Minor Salivary Glands*

Briefly, SG biopsies (~5 per patient) were immediately placed on ice. SGs were then gently transferred to a sterile 100 mm tissue culture dish and minced into small pieces (< 1 mm) using a disposable scalpel. Dissociation of lobules was then performed for 40 mins in heated sleeves at 37 °C in an OctoMACS tissue disruptor using the Miltenyi Multi-tissue Dissociation Kit A and Multi_A01 in C-type tubes using the tumor dissociation kit 1 enzyme set. Complete dissociation of samples was verified through visual inspection prior to filtration. Tissue samples were then filtered through a 70 - 30 µm MACS SmartStrainer and rinsed with 1X HBSS ($Ca^{2+}$ and $Mg^{2+}$ free) supplemented with 0.4% ultra-pure, DNase/RNase-free BSA. Using 1X HBSS, the solution was then washed and pelleted twice at 300 g for 10 mins at ambient temperature. Cells and viability were quantified using a hemacytometer after trypan blue staining; 100,000 cells were used for capture and single cell sequencing.

*scRNAseq Processing & Analysis*

SG biopsies (experiments, n=2; control, n=6) were obtained and processed for scRNAseq analyses as described previously (**Figure 2A**) and analyzed using Seurat v.5 (R Version 4.3.2).[2] The SoupX package[3] was utilized to bioinformatically remove ambient mRNA contamination from all eight patient samples, followed by doublet discrimination using the scDblFinder function. Quality control (QC) was independently performed on individual samples to remove cells with elevated mitochondrial and ribosomal content as well as cells with less than 200 genes. The SoupXcorrected filtered gene barcode matrices were then merged and normalized using Seurat's NormalizeData() workflow. Integration was performed using the STACAS package.[4] The top 10 most variable genes were determined using Seurat's FindVariableFeatures(), and the data was scaled with ScaleData(). Principle components analysis (PCA) was performed using RunPCA(), and the number of principal components (PC) was determined via dimensionality analyses using an ElbowPlot. From this, 20 dimensions were chosen, and Uniform Manifold Approximation

Projection (UMAP) was carried out using Seurat's RunUMAP() for visualization and non-linear dimension reduction.

Cell clusters were manually annotated, and the cluster for red blood cells was removed. A total of 34,694 genes among 27,355 cells (20,614 and 6,741 cells from the healthy and ICI-GS, respectively) remained for final clustering and annotation (**Figure 2A, Supplemental Figure 1C**). Subclustering was subsequently performed to better distinguish between cell types, and the proportion of each cell type across each condition was visualized (**Figure 2B**). Mucous (*MUC5B* and *WFDC2*) and seromucous acini (*PRR4* and *AQP5*) comprised the acinar populations. Myoepithelial cells (*ACTA2* and *KRT14*), pericytes (*RGS5* and *PDGRFB*), endothelial cells (*RAMP2* and *AQP1*), and fibroblasts (*CXCL14* and *DCN*) comprised the connective tissue. Subclustering of the data revealed 7 unique immune cell types: B cells/plasma cells (*CD79A),* $CD4^+$ T cells (*CD3E*, *CD4, CD40LG*), $CD8^+$ T cells (*CD3E*, *CD8A,* and *CD8B*), proliferating T cells *(CD3E, MKI67),* M1 macrophages (*IL1B*), M2 macrophages (*MRC1*), and monocytes (*NR4A1*).

Importantly, there were cells that co-expressed genes typical of both seromucous cells (*PRR4* and *WFDC2*) and immune markers (*HLA-DRA*, *HLA-DRB1*, and *CD40*). These cells were posited to be immune-engaged seromucous (IE-seromucous) cells and were only present within the ICI-GS biopsy. Strikingly, HLA-DRA was present in ~96% of IE-seromucous cells but <3% of healthy seromucous cells. Similarly, IE-mucous acinar cells showed high expression of mucous markers (*MUC5B*) and immune markers (*HLA-A* (~92%), *HLA-B* (~97%), and *CD74* (~89%)) and were nearly entirely absent from HV glands. The top three variably expressed genes per cluster were analyzed using the "MAST" method and are provided in **Supplemental Figure 2B**.[5]

Differentially expressed markers between conditions were similarly identified using the "MAST" method and visualized with volcano plots using Qiagen's Ingenuity Pathway Analysis (IPA).[6] These genes were then analyzed for pathway enrichment using IPA. A heatmap of the conserved pathways among the different clinical conditions and cell types was created, keeping pathways that had less than five *z*-scores equal to zero among the conditions plotted for visualization (**Figure 2C**). Additionally, ligand-receptor interactions were examined in the single cell object using the Seurat's CellChat() workflow (**Figures 2E**). To estimate senescence, the human Universal Senescence Index (hUSI) was applied to the scRNAseq data (Wang, et al., 2025).

*Spatial Transcriptomics Processing and Analyses*

Xenium is a spatial transcriptomics tool from 10x Genomics that allows for the visualization of genes by their location within the tissue. This platform enabled us to study the cell-to-cell communications, spatial localization patterns, and cellular microenvironments. Formalin-fixed, paraffin-embedded (FFPE) tissue sections from HVs, ICI-GS-pre, and ICI-GS-post were processed following the protocol provided by 10x Genomics (Xenium *In-Situ* Gene Expression); a commercially available 280-plex breast cancer panel was used.

Before normalizing the data with Seurat's NormalizeData() workflow, QC was performed on the spatial transcriptomics (ST) data, removing cells with less than 20 molecules and 10 genes. Briefly, spatially informed expression data from 277,511 cells were returned (143,200 and 134,311 from HV and ICI-GS, respectively). The FindTransferAnchors() workflow was used to transfer cell type labels from scRNAseq ('reference dataset') to Xenium ('query dataset') with high confidence (~95%) (**Figure 2A**).[8] In total, 14 unique cell types were annotated in the spatial analyses. Briefly, the top 10 most highly variable genes were determined using FindVariableFeatures(), before scaling the data with Seurat's ScaleData() workflow. PCA were computed with RunPCA(), and PC (20) were determined with the ElbowPlot(). UMAPs were created using 20 dimensions within the RunUMAP() workflow for health and disease. The TransferAnchors() workflow was used between the annotated scRNAseq and ST datasets,[8] with 275 genes conserved among the datasets. The proportion of cells associated with each cluster was plotted (**Supplemental Figure 1C**), and a scatter plot between the cell proportions in single cell and spatial data revealed a strong, significant correlation (Corr. , $p < 0.005$; *data not shown*). To visualize the spatial plots and the cell mask with identical cluster annotations, the data were uploaded onto Xenium Explorer (version 2.0). A full spatial plot of each of the tissues for each condition is provided in **Supplemental Figure 1D**, and H&E stains were overlaid with gene markers from spatial transcriptomics data. In particular, the green, red, and yellow dots represent composite epithelial (*AQP3*, *CDH1*, *EGFR*, *EPCAM*, *FOXA1*, *KLF5*, *KRT5*, *KRT8*, *MLPH*, and *TACSTD2*), T cell (*CCL5*, *CD3E*, *CD3G*, *CD27*, *CD4*, *CD8A*, *GZMA*, *GZMK*, *IL2RG*, *IL7R*, *NKG7*, *SLAMF7*, and *TRAC*), and macrophage (*AIF1*, *CD68*, *CXCL16*, *CXCL5*, *CX3CR1*, *C1QC*, *FCER1G*, *FCGR3A*, *IGSF6*, *ITGAX*, *MNDA*, *MRC1*) gene markers, respectively (**Figure 2H**). To understand the pathogenic niches and cellular interactions between cell types in health and disease before and after prednisone therapy, closeness centrality (indicator of centrality of cell subset), degree centrality (indicator of connections for a cell subset), average clustering (indicator of overall connectedness of a cell subset) (**Figure 2J**, **Supplemental Figures 1E**) were conducted using Python 3 on JupyterLab.
